# Randomized incentives to increase participation in COVID testing in rural Kenya

**DOI:** 10.64898/2026.02.15.26346122

**Authors:** Benard Chieng, Yoshika S. Crider, Colin Aitken, Sylvie B. Araka, Richelle W. Kihoro, Henry Kanyi, Julie E. Powers, Brandon Tan, Abigail P. Paulos, Alexandre Simoes Gomes, Carol Nekesa, Cecilia Nekesa, Blastus Bwire, Dennis Odhiambo Allela, John Kiiru, Michael Kremer, Sammy M. Njenga, Amy J. Pickering

**Affiliations:** Kenya Medical Research Institute, Nairobi, Kenya; Division of Environmental Health Sciences, University of Minnesota School of Public Health; King Center on Global Development, Stanford University; Civil & Environmental Engineering, UC Berkeley; King Center on Global Development, Stanford University; Civil and Environmental Engineering, University of California, Berkeley; Development Innovation Lab, University of Chicago; Harvard University; Remit Global Research Center; University of Chicago, Chicago, Illinois; Civil and Environmental Engineering, University of California, Berkeley Chan-Zuckerberg San Francisco Biohub

**Keywords:** COVID-19 testing, non-response bias, incentives, surveillance

## Abstract

Low participation in public health testing can lead to biased estimates of disease prevalence and inefficient allocation of public health resources. We evaluated the impact of monetary incentives and revisits on participation in door-to-door COVID testing in rural western Kenya. We conducted a cross-sectional study of all residents in 12 villages in rural western Kenya. We offered an incentive of KSh 200 (1.85 USD), KSh 350 (3.23 USD), or KSh 700 (6.47 USD), randomized at the household level, for each household member >3 months old who participated in COVID testing. Among 7,049 individuals, 5,659 individuals consented to testing. Overall, after revisits, 78.2% consented in the KSh 200 group, 80.6% in the KSh 350 group, and 81.9% in the KSh 700 group. Among individuals offered KSh 200, revisiting increased the consent rate from 68.1% to 78.2%, or 7.3 percentage points higher than the first visit consent rate in the KSh 700 group. Participation was very high (96.9%) among available individuals, but 17.2% of individuals were never available. Offering KSh 200 minimized the cost per consenting individual in our sample, even with revisits, compared to higher amounts. However, individuals in the KSh 700 group were slightly more likely to test positive, which suggests these individuals are missed at lower incentive amounts. Overall 0.3% (95% CI: 0.2, 0.5) tested positive for current infection by qPCR on nasal swabs, and 8.6% (7.7, 9.6) tested positive for SARS-CoV-2-specific antibodies by ELISA on dried blood spots. Accounting for nonresponse bias suggests this COVID population burden may be an underestimate. Our findings suggest incentives can increase participation in household door-to-door public health surveillance testing and may improve disease prevalence estimates by reducing nonresponse bias. However, high incentives may not be cost effective when low incentives motivate very high levels of participation.

**Significance Statement:** Achieving high levels of participation in testing can inform public health strategies for reducing the spread of infectious diseases. Cash incentives are one tool for increasing participation, but there is little evidence on the effectiveness of this approach in low-resource settings. We find that cash incentives increase participation in free COVID-19 testing in rural Kenya and may reduce nonresponse bias.

## Introduction

Testing to identify and treat positive cases is a key public health strategy for controlling infectious diseases. However, low rates of testing coverage and participation can contribute to biased estimates of disease prevalence and inadequate information to inform public health investment and action. (1, 2) This was clearly demonstrated during the COVID-19 pandemic, where low rates of testing led to significant underestimation of disease burden.(1) Even where cost was not a barrier, low participation was observed. For example, in Luxembourg, less than half of those invited took part in free testing at testing sites.(3) In northern California, USA, participation rates for free door-to-door testing for COVID-19 were less than 51%.(4)

Monetary incentives are one strategy to increase participation. These have been used across high- and low-income settings to encourage pro-public health behaviors, for example, to increase response rates for mailed health access surveys (e.g., Medicaid surveys in the United States (5)), increase utilization of infectious disease testing (e.g., HIV/STIs (6, 7)), increase vaccine uptake (e.g., COVID-19 vaccines in Sweden (8), tetanus vaccines in Nigeria (9)) and improve treatment adherence (e.g., tuberculosis treatment in Swaziland (10)). Research on health promotion suggests that cash payments, particularly when guaranteed and immediately delivered, are likely to be more effective incentive strategies than non-cash or lottery-based incentives.(11) However, there is little evidence on their cost-effectiveness, especially in low-resource settings, which is a key question for policymakers.

We fill this evidence gap by examining the effect of monetary incentives on testing participation in the context of COVID-19 testing in Kenya. The country reported its first SARS-CoV-2 infection on March 13, 2020. Soon after, the Government of Kenya closed schools and colleges, restricted foreign travel, and limited gatherings to prevent the spread of COVID. However, sparse testing in rural counties meant there was limited information about the spread of the disease across much of the country.(12) Here, we evaluate the impact of randomized, conditional cash incentives on availability and consent to participate in free, door-to-door testing for SARS-CoV-2 infection in rural Western Kenya. Our aims were to: (1) assess the effect of cash incentives on participation in COVID testing, (2) assess the cost-effectiveness of incentives and household revisits to increase participation rates, and (3) identify evidence of nonresponse bias.

## Results

From 12 February 2021 through 25 April 2021, we recorded 7,049 household members in 1,345 households and enrolled 5,659 individuals in 1,326 households (Figure 1) across 12 villages. Our sample included 2,291 adults and 3,368 minors. On average, 4.3 individuals were enrolled per household. Among those who participated, the median age was 14 years (range: 3 months to 97 years) and 44% were male.

**Figure 1.**
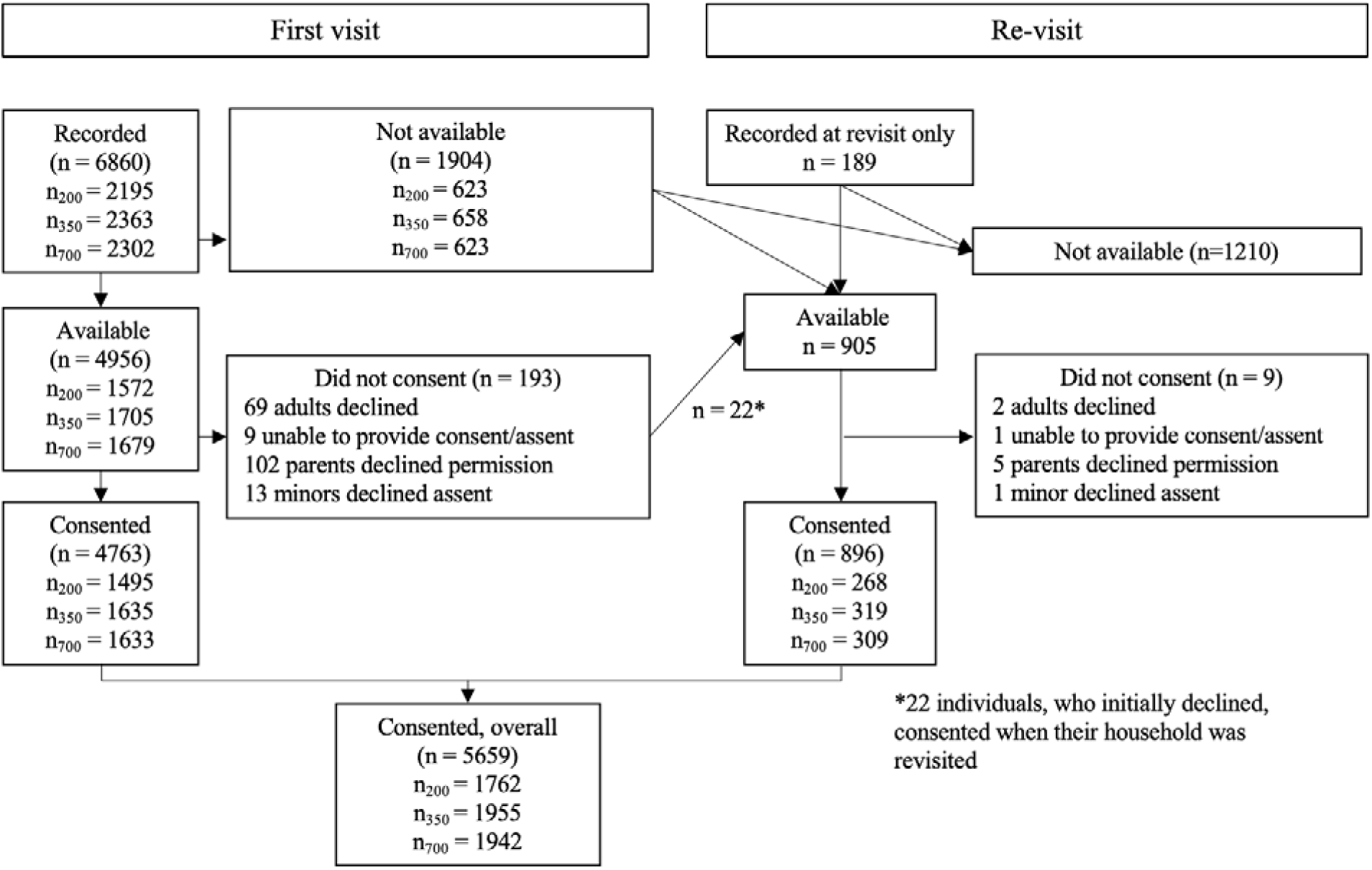
Study flow chart *Gift amounts and study participation*

The proportion of available individuals consenting was high for all gift amounts across first visits (95.1-97.3%), revisits (98.5-99.7%), and all visits (96.0-98.2%) (Table 1). In the first visit, increasing the gift amount from Kenyan shillings (KSh) 200 to 700 increased overall consent by 3.8 percentage points (5.6% increase). This was driven by both an increase in availability (2.2 p.p. / 3.1%) and consent among those available (2.4 p.p. / 2.5%), among individuals offered KSh 700 versus 200. Although effects were also positive for KSh 350 versus 200, they were not statistically significant at the p=0.05 threshold (Table 2).

**Table 1.**
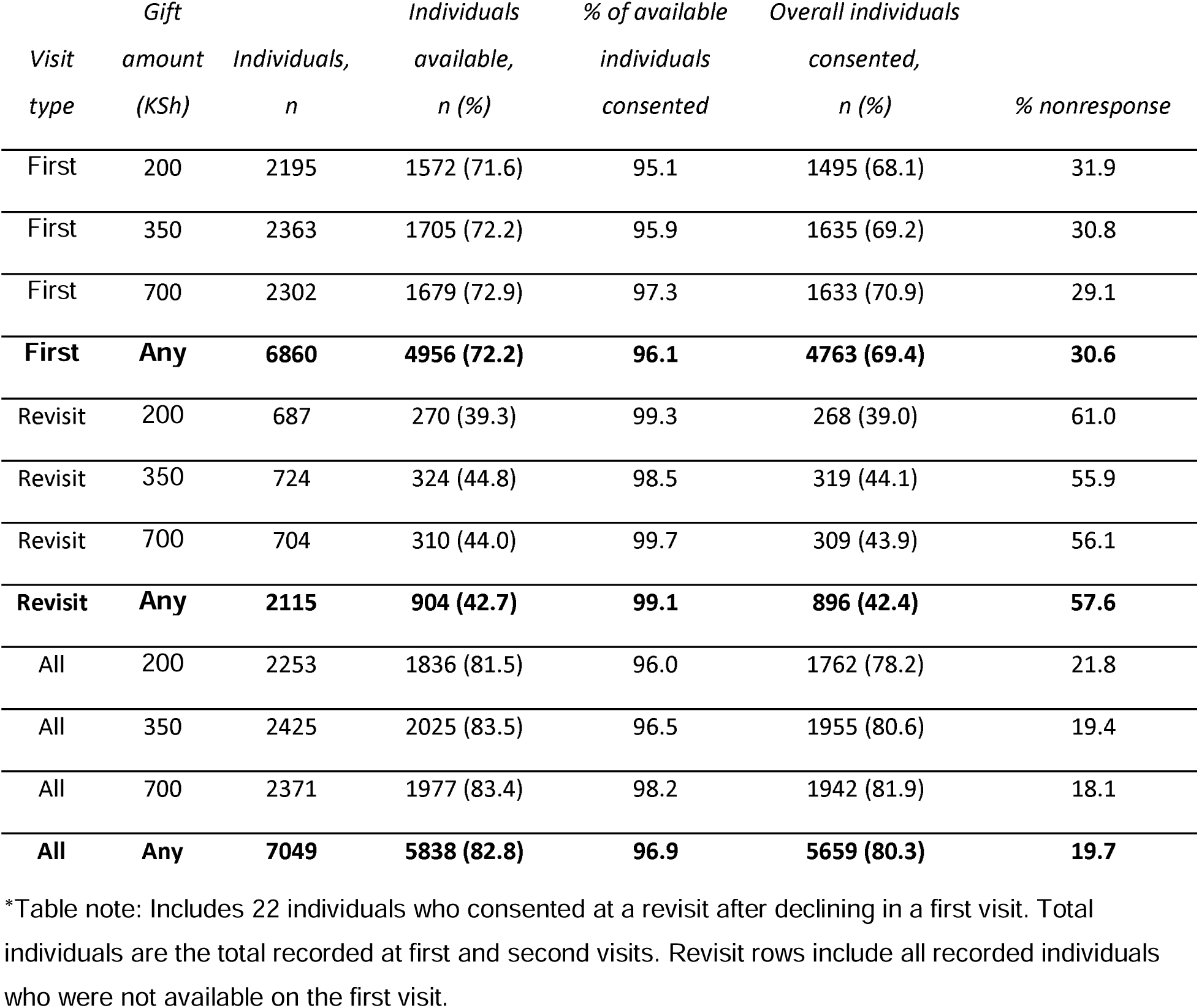
Availability and consent first visits, revisits, and overall.

**Table 2.** Regression results, first visit.

|  | *Dependent variable:* |  |  |
| --- | --- | --- | --- |
|  | Consented<br>(1) | Available<br>(2) | Consented \| Available<br>(3) |
| Gift = 350 | 0.021<br>(0.018) | 0.012<br>(0.017) | 0.013<br>(0.009) |
| Gift = 700 | 0.038**<br>(0.018) | 0.022<br>(0.017) | 0.024***<br>(0.009) |
| Mean 200KSH | 0.68 | 0.72 | 0.95 |
| Village fixed-effects | Yes | Yes | Yes |
| Enumerator fixed-effects | Yes | Yes | Yes |
| Observations | 6,860 | 6,860 | 4,956 |
| R<sup>2</sup> | 0.040 | 0.046 | 0.032 |
| Adjusted R<sup>2</sup> | 0.035 | 0.041 | 0.026 |
| Residual Std. Error | 0.452 (df = 6828) | 0.438 (df = 6828) | 0.191 (df = 4924) |
| *Note:* | | * $p<0.1$; ** $p<0.05$; *** $p<0.01$ | |

Revisits targeted those unavailable during first visits (n=1904) and additionally included individuals first recorded during a household revisit (n=189). At revisits, 42.7% of target individuals were available, of whom 99.1% consented. Revisit consents include 22 individuals who declined during a first visit, but subsequently consented during a revisit. The overall revisit consent rate was 39.0% among individuals who were offered KSh 200, 44.1% for 350, and 43.9% for 700.

Across all visits, the overall consent rate was 78.2% among individuals who were offered KSh 200, 80.6% for 350, and 81.9% for 700 (Table 1). Increasing the gift incentive from KSh 200 to 700 increased consents by 4.3 p.p. (5.5%), by increasing availability by 2.5 p.p. (3.1%) and consent among available individuals by 2.4 p.p. (2.5%). Increasing from KSh 200 to 350 had a positive, but not statistically significant, effect on consent and availability at the p=0.05 threshold (Table 3). There was no evidence that the consent rate changed as the overall share of a village surveyed increased (SI Table 1).

**Table 3.** Regression results, all visits.

|  | *Dependent variable:* |  |  |
| --- | --- | --- | --- |
|  | Consented<br>(1) | Available<br>(2) | Consent \| Available<br>(3) |
| Gift = 350 | 0.032*<br>(0.016) | 0.025<br>(0.016) | 0.010<br>(0.008) |
| Gift = 700 | 0.043***<br>(0.016) | 0.025<br>(0.015) | 0.024***<br>(0.007) |
| Mean 200KSH | 0.78 | 0.81 | 0.96 |
| Village fixed-effects | Yes | Yes | Yes |
| Enumerator fixed-effects | Yes | Yes | Yes |
| Observations | 7,049 | 7,049 | 5,838 |
| R<sup>2</sup> | 0.043 | 0.050 | 0.028 |
| Adjusted R<sup>2</sup> | 0.038 | 0.045 | 0.022 |
| Residual Std. Error | 0.390 (df = 7016) | 0.369 (df = 7016) | 0.170 (df = 5805) |
| *Note:* |  | * p<0.1; ** p<0.05; *** p<0.01 |  |

We observed differential effects of incentives stratified by county, with the effect primarily due to responses in Vihiga County. Overall availability was higher in Vihiga County (80%, versus 70% in Bungoma County), and a gift incentive of KSh 700 in Vihiga County increased consent among available individuals by 6.1 p.p. (6.7%), and overall consent by 8.7 p.p. (11.9%) (<u>SI</u> Table 2). In contrast, no statistically significant effects of gift incentives were observed in Bungoma County alone.

### Revisit costs

Overall, 15.8% of consents were obtained during revisits. We estimate that a single first visit (per individual) cost KSh 630 (∼6 USD) and a second visit cost 2x a first visit cost (based on actual monthly study costs and total visits conducted, higher costs resulted from additional transport necessary to reach revisit households spread over a larger geographic area than first visits), plus the incentive amount upon consent. Based on observed consent rates at first visit and revisits, we estimate the marginal cost of an additional first visit consent to be KSh 9581 when increasing the incentive from KSh 200 to KSh 350 and 14928 when increasing the incentive from KSh 350 to KSh 700, and the marginal cost of an additional overall consent to range from 5219 when increasing the incentive from KSh 200 to KSh 350 and 21418 when increasing the incentive from KSh 350 to KSh 700 (Table 3). Across fixed incentive amounts, the marginal cost of additional revisit consents is similar (KSh 3208-3572).

**Table 3.** Marginal cost per consent. The historical exchange rate was 1 USD: KSh 108 in May 2021.

| Gift amount<br>(Ksh) | Marginal cost per<br>consent, first visits<br>only, compared to<br>next lower incentive<br>(Ksh) | Marginal cost per<br>revisit consent at<br>fixed incentive<br>amounts (KSh) | Marginal cost per<br>consent, overall,<br>compared to next<br>lower incentive (Ksh) |
| --- | --- | --- | --- |
| 200 | 1125* | 3430 | 1448** |
| 350 | 9581 | 3208 | 5219 |
| 700 | 14928 | 3572 | 21418 |

\*Cost per consent based on incentives paid and visit costs for 7049 first visits
\*\*Cost per consent based on incentives paid and visits costs for 7049 first visits and 2002 revisits. Higher incentives had reduced revisit costs because of slightly higher availability at first visit.

### SARS-CoV-2 infection prevalence and symptoms

We obtained results from ELISA and qPCR tests for 5423 and 5617 individuals, respectively. Overall 0.3% (95% CI: 0.2, 0.5) tested positive by qPCR, and 8.6% (7.7, 9.6) tested positive for SARS-CoV-2-specific antibodies by ELISA. Cumulative incidence of infection was 9.0% (8.1, 10.0). Stratified by age, seropositivity was highest among 18-60 year olds (11.6% (10.2, 13.1)) and lowest among 0-5 year olds (4.0% (2.8, 5.3)). Prevalence of current infection was highest among >60 year olds (1.1% (0.3, 2.3)) and lowest among 0-5 year olds (0.1% (0.0, 0.3)) and 6-17 year olds (0.2% (0.0, 0.4)) (Figure 2, <u>SI</u> Table 6). There was no difference in infection results by sex. Stratified by county, prevalence of current infection and seropositivity were more than double in Vihiga County (0.6% (0.2, 1.2) and 16.0% (13.3, 18.7)) compared to Bungoma County (0.3% (0.1, 0.4) and 6.9% (6.0, 7.8)) (<u>SI</u> Figure 2, Tables 6-8).

### Factors associated with infection and consent

We found no association between infection status and self-reported illness, which was defined as fever, cough, and/or difficulty breathing since government school closures due to COVID began in March 2020 (4% among negatives versus 4% among positives, p=0.61)). Of the 236 respondents who self-reported illness, 85 visited a hospital or clinic due to their symptoms.

Individuals were diagnosed by a health professional with malaria (53/85), cold or flu (20/85), or respiratory infection (11/85). 19 individuals suspected their symptoms may have been caused by COVID-19, but none were diagnosed with COVID-19. Self-reported symptoms in the prior 7 days that were more commonly reported (p<0.05) among individuals with positive qPCR (n=19) tests versus negative qPCR tests included chills (16% vs 4%), unusual muscle pains (16% vs 3%), unusual fatigue (11% vs 3%), loss of appetite (16% vs 2%), repeated shaking with chills (21% vs 2%), and unusual fatigue (11% vs 3%) (<u>SI</u> Table 9).

There was no difference in self-reported financial stress across incentive amounts, based on questions about whether the (adult) participant or a household member had been forced to borrow money or sell assets in the prior 30 days. Living in a household with more than 6 people was also not associated with consent across incentive amounts (SI Figure 1).

### Potential non-response bias and infection estimates

We observed COVID-19 rates among people who consented to be tested, but these rates were subject to nonresponse bias: if people who were not tested had systematically different COVID rates than people who were, the infection rates we measured would not be reflective of the population as a whole. After including week fixed effects to account for rising infection rates over time, individuals who were reached on return visits did not have significantly higher COVID-19 infection rates than their counterparts (Table 4).

**Table 4.** Infection rates by visit type.

| **Infection Rates by Visit Type** |  |  |
| --- | --- | --- |
|  | *Dependent variable:* |  |
|  | Any Positive COVID Test |  |
|  | (1) | (2) |
| Return Visit | 0.021<sup>*</sup><br>(0.011) | 0.002<br>(0.011) |
| Constant | 0.087<sup>***</sup><br>(0.004) |  |
| Week Fixed Effects | No | Yes |
| Observations | 5,386 | 5,386 |
| R<sup>2</sup> | 0.001 | 0.028 |
| Adjusted R<sup>2</sup> | 0.0005 | 0.026 |
| Residual Std. Error | 0.286 (df = 5384) | 0.282 (df = 5374) |
| *Note:* | <sup>*</sup>p<0.1; <sup>**</sup>p<0.05; <sup>***</sup>p<0.01 |  |

We find evidence of non-response bias comparing people who consented to testing when offered higher compensation, with COVID rates in the KSh 700 group roughly 2.5 percentage points higher than in the KSh 200 group (Table 5).

**Table 5:**
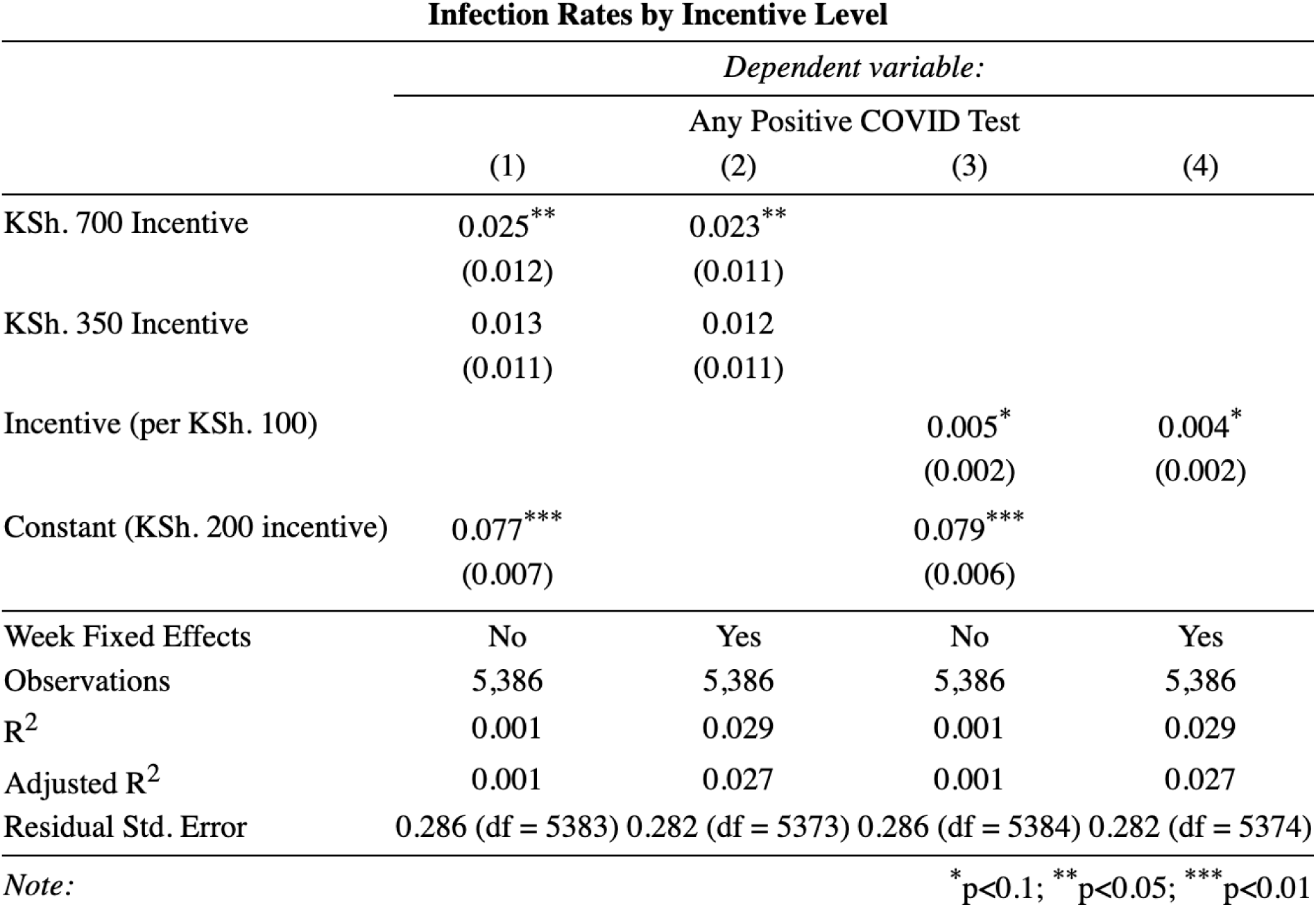
Results of regressing COVID seroprevalence on compensation levels, either as a categorical variable (columns 1 and 2) or a continuous variable (columns 3 and 4). All standard errors clustered at the household level.

## Discussion

We found high levels of participation in free, door-to-door COVID testing in rural Kenya in exchange for cash incentives of KSh 200-700. Consent was mostly limited by availability, as 95% or more of available individuals consented across all gift amounts. Increasing the incentive from KSh 200 to 700 had a positive effect on consent among available individuals, but these additional consents were expensive to obtain.

The overall 80.3% participation rate was lower than COVID testing campaigns in China,(13) similar to Slovakia,(14) and higher than Luxembourg(3) and the United States.(4) We consider several factors that may have inclined individuals to participate, in addition to the incentive amounts that were perhaps high for the area. First, familiarity with the model of community health workers (CHWs) conducting home visits may have contributed to high participation. For example, a door-to-door HIV testing campaign conducted over a decade ago in Western Kenya had a nearly 90% consent rate among eligible adults, establishing that a home-based testing approach can be successful in this setting.(15) Social norms may also have encouraged participation in this community-wide public health campaign; some evidence indicates that psychological motivators are more effective than monetary incentives outside of Western, high-income cultures.(16) Concern about risk of infection was low and therefore unlikely to have influenced high participation – approximately one-fifth of adults reported feeling that they or their household was at risk of contracting COVID. The majority of individuals who reported visiting a clinic for symptoms commonly associated with COVID were subsequently diagnosed with malaria, and some of the self-reported symptoms that were more prevalent among current COVID-19 cases in this sample are common symptoms of malaria. Bungoma and Vihiga counties are located in Kenya’s lake-endemic region, where, in 2015, approximately a quarter of children under 14 years were positive for malaria parasites by blood microscopy.(17) Although national guidelines recommend seeking care within 24 hours of symptoms, a study in western Kenya found that less than 20% of individuals with febrile illness sought care at a health facility.(18)

We found that increased incentives were not cost-effective in our sample due to very high participation at the lowest offered incentive. Revisits with KSh 200 result in a higher overall consent rate at a lower marginal cost per consent than first visits with KSh 700. This also suggests there may be an even smaller incentive that could be optimally cost effective. We do not know whether a zero incentive would have resulted in high participation in our sample, though evidence from elsewhere suggests some non-zero incentive is important. For example, in rural Malawi, a small incentive doubled the likelihood that individuals picked up HIV test results from counseling centers following free, door-to-door testing, compared to no incentive.(19) It would be informative for future studies of randomized incentives to include a smaller or zero incentive group for reference. Availability was not impacted by incentive amount and, while even higher incentives could be investigated, the risk of coercion, another criticism of financial incentives (20), would increase. One reason participants may not have been available even for revisits is that the study period overlapped with harvesting and sowing seasons for key crops in the region.(21) Allowing for alternative methods of gathering survey data (e.g., through phone calls or text surveys), at-home testing or self-sampling, early morning and night-time visits, or testing at the workplace may have been necessary to reach these individuals.

For context, our overall estimates of cumulative infection (11%) in rural Bungoma and Vihiga counties from February through April 2021 varied compared to estimates elsewhere in Kenya. By the end of April 2021, there had been approximately 160,000 confirmed cases of COVID-19 in Kenya.(22) In Kenya, COVID-19 cases first peaked in August 2020 followed by a second and third wave in November 2020 and March 2021, respectively (23). Our study coincided with a third wave. Earlier studies conducted in an urban region in Nairobi County between November to December 2020 found a seroprevalence of 33.0-43.4%, several times higher than we found, detected through ELISA.(24, 25) A similarly timed study conducting household visits at health and demographic surveillance sites in Kisumu, Nairobi, and Kilifi from Dec 2020-May 2021 found higher seroprevalences than our study ranging from 25-50% (26, 27), though our sites are more rural. Another study conducted in Coastal Kenya with an overlapping time period to ours found an almost seven times higher prevalence of 7.7% detected through qPCR.(26) Later studies carried out among individuals seeking COVID-19 tests at hospitals Kisumu and Siaya counties in Western Kenya, between December 2021 and February 2022, showed an overall seroprevalence of 76.5% and 29.6% for IgG and IgM respectively and 33.7% had a positive PCR test (28). Our study yielded lower seroprevalence estimates compared to those obtained from essential workers such as among health care workers (20.8%) between July to December 2020 (29) and truck drivers (42.3%) in October 2020 (30), and among blood donors (48.5%) between January to March 2021.(31)

We did not find evidence that COVID rates were higher on revisits than initial visits in our sample, but we did find evidence that COVID rates were higher for individuals who would only consent to test when offered higher compensation. This implies that nonresponse bias would have been present had we not offered higher incentives and creates a concern that it might still be present among people we were not able to reach. In a study of incentives for COVID-19 testing and vaccination in the United States, researchers found that low incentives reduced self-reported intentions for vaccines but not for testing, highlighting the importance of identifying the correct incentive amount.(32) In rural Kenya, it seems unlikely that lower incentive amounts were a disincentive but rather it did not balance the opportunity cost for some participants of the time or discomfort required to participate.

Our study has several strengths. First, our free, door-to-door testing approach resulted in estimates of COVID prevalence that are likely representative of community burden. Neither symptoms nor seeking medical care were associated with infection outcomes, indicating that testing only symptomatic individuals who sought medical care would have resulted in biased estimates of community prevalence. Additionally, because there are no costs to participants (e.g., for the test itself or transportation to testing sites), our sample includes community members for whom those costs may otherwise be a barrier. Instead, reasons to decline participation could include the opportunity cost of the time to participate (e.g., missed wages), hesitancy to share information, or reluctance to provide biological samples. Second, by including both antigen (qPCR) and antibody (ELISA) testing, we are able to assess both a snapshot of transmission risk and an estimate of the cumulative incidence of infection.

There are important limitations to our data as well. First, we have no zero-incentive reference group. Our findings at KSh 200 suggest small incentives in this population can lead to high participation, but we lack data to know whether 0 incentive may actually have a similar result. We do not have data on individuals who declined to participate, so we cannot determine if or how non-participants differ from participants. Our cross-sectional study design has only one time point, so we cannot infer any directionality of SARS-CoV-2 transmission through households and communities. Symptoms and behaviors were self-reported and thus subject to courtesy and recall bias, although for most questions we limited recall to the prior 7 days to reduce the latter bias. We may be underestimating cumulative incidence of infection due to waning seropositivity. Our study began 339 days after the first official case was recorded in Kenya, though a similar-performing immunoassay was shown to reliably detect antibodies at least 8 months post-infection.(33, 34) Additionally, due to problems with global supply chains during the pandemic, we were unable to procure enough ELISA kits to analyze all of our samples. However, this affected less than 5% of collected samples. Finally, we had imperfect randomization in the last few weeks of the study, when some enumerators appeared to restart surveys until the highest incentive value was randomly generated; we therefore restricted our sample to exclude data collected in the final villages. Ensuring enumerator adherence to study protocols may pose a challenge for future implementation of randomized incentives; it will be important to consider strategies to mitigate this.

## Conclusion

Approximately one year after the first case of COVID-19 was reported in Kenya, participation in free door-to-door COVID testing was very high among available individuals in exchange for cash incentives ranging from KSh 200-700 per person. A gift amount of KSh 700 increased consent at first visits compared to the lowest incentive amount, but the consent rate was limited by availability, and revisits had the greatest overall effect. Overall, approximately 15% of individuals remained unavailable, limiting community consent rates. Appropriately valued cash incentives may be a cost-effective strategy to increase household data collection efficiency, especially when revisit costs are high, but maximizing participation will require strategies to increase availability. These may be individuals who are at increased risk for COVID-19 due to their occupations, and, because we observed higher test positivity in the highest incentive group, we estimate that non-response biased our estimates of community COVID prevalence slightly downward. In a future paper, we will address the problem of nonresponse bias more directly by considering a researcher trying to minimize uncertainty from sampling variation and nonresponse bias given a fixed budget and solving for optimal behavior under different assumptions. To meaningfully increase participation in community-wide testing and avoid non-response bias, it may be useful to combine incentives with alternative methods of participation such as phone-based surveys, at-home testing or self-sampling, and testing options at the workplace.

## Methods

### Study design

We conducted a cross-sectional population-based study in rural villages in Vihiga and Bungoma Counties in Kenya’s Western region from February 12 through April 25, 2021. These villages were participating in an ongoing impact evaluation of a water treatment intervention on child survival, which allowed us to take advantage of the resources and expertise of a data collection team already working in the study area. Before embarking on this study, the research team engaged with Kenya’s Ministry of Health at the National, County, sub-county and ward levels. Community Health Promoters (CHPs) were recruited to assist in the introduction of the study in each village. Prior to visiting each village, a meeting was organized with village elders and other key opinion leaders. This meeting introduced the study objectives and activities.

### Participants

Eligible individuals included anyone over the age of 3 months residing in 15 villages within our study area. Drawing on village census information collected in a previous study; we conducted random sampling from geographic areas within a feasible distance from our field lab to select enough villages to enroll 3000 households. Ten villages were from Bungoma County, and 5 villages were from Vihiga County. At each household, enumerators knocked on doors and asked to speak with the female head of household or, if none were available, another adult household member. Written consent was obtained from all adults (18+ years old) and mature minors (<18 years old and married, pregnant, a mother, or household head; hereafter referred to as adults); parent/guardian permission was obtained for all minors (<18 and not mature minor), and verbal assent was additionally obtained from all minors 7+ years of age. Not all household members were required to participate in order for others to enroll.

### Gift incentives

Each household was randomly assigned to an M-Pesa mobile money transfer of KSh 200 (1.85 USD), KSh 350 (3.23 USD), or KSh 700 (6.47 USD) per person within the household who consented to SARS-CoV-2 testing (both a nasal swab and dried blood spot collection). The highest amount was selected to compensate for approximately one day of missed wages in the study area. At households where no one was home, no individuals were recorded, and enumerators returned on another day. At households where at least one adult was home, the names of all eligible household members were recorded as potential participants, and enumerators scheduled a return visit at a mutually convenient time. The “first visit” was defined as the first visit for which an adult was home. The household’s randomized gift amount was assigned at the first visit. Individuals who were available but did not consent to participate at the first visit were allowed to change their minds and participate if their household was subsequently revisited.

### Household survey

A survey was administered to each adult household member by the study team. Data were collected about knowledge of COVID-19 symptoms, self-reported COVID-19 symptoms experienced since the start of the pandemic in Kenya (March 2020) and specific symptoms in the prior 7 days, risk perceptions, potential behavioral risk factors for infection (e.g., number of non-household contacts and mask wearing), and health-care seeking behavior. We collected symptom data for minors under 5 years of age from their parent/guardian.

### Sample collection and processing

Trained laboratory technicians collected nasal swab samples and dried blood spots from each participant. Swabs were shortened by sterilized scissors and placed into 2 ml microcentrifuge tubes. Dried blood spots (DBS) were collected via finger prick; a participant’s finger was cleaned with a sterile alcohol swab and pricked using a disposable lancet; blood was collected on Whatman filter paper. Samples were shipped on dry ice to a KEMRI laboratory in Nairobi for analysis. Individuals with positive qPCR results were informed via phone call.

### Detection of SARS-CoV-2 using molecular techniques

Testing was first done on pooled samples to reduce the number of RT-qPCR tests needed. To each individual sample, we added 1.3 ml 1X PBS and vortexed for 15 seconds prior to pooling. To prepare a single pooled sample, 35 microlitres were selected from each of 5 samples and combined in a 1.5 ml microcentrifuge tube. RNA was extracted using QIAmp Viral RNA mini kit (QIAGEN, Hilden, Germany) and eluted using 60 microlitre of nuclease free water. RT-qPCR assays were performed using a StepOnePlus™ real time PCR instrument. Reactions were run on 96-well plates with a reaction volume of 20 microlitres. 3 assays (N1, N2 and RNAse P) were run on each plate to test for SARS CoV-2. Samples negative in pooled testing were reported as negative, whereas samples positive in pool testing were individually re-extracted and re-tested to identify the individual positive sample(s).

### Detection of SARS-CoV-2 using enzyme-linked immunosorbent assay (ELISA) (antibody detection)

We used the commercial Wantai SARS-CoV-2 Ab ELISA kit (Wantai Biological Pharmacy Enterprise Ltd, Beijing, China) to analyze dried blood spots (DBS) samples for SARS-CoV-2–specific antibodies (IgG, IgM) with a slight modification to the manufacturer’s instructions. To rule out cross-reactivity with antibodies to other pathogens (35), we first validated the ELISA test kit using pre-pandemic DBS samples (n=90) collected in a coastal region of Kenya in 2015. With these samples, an initial validation assay adhering to manufacturer’s instructions resulted in a high number of false positives. Subsequently, we adopted procedures detailed in Ngere et al.(25), which eliminated false positives. Briefly, a series of optimization tests demonstrated that 10 washes were necessary to minimize background cross-reactivity in the assay, as opposed to the 5 washes recommended by the manufacturer. See SI for further protocol details. Due to supply chain limitations, we were unable to procure enough ELISA kits to process all DBS samples (4.8% (n=370) were not processed).

### Statistical analysis

A prespecified analysis plan is available at https://osf.io/rsyeg. We cleaned data in Stata (MP version 17) and performed statistical analysis with RStudio version 4.3.1. Our analytic dataset excludes the last 3 villages visited in the study. In these villages, enumerators admitted to restarting surveys to provide higher incentives for households, violating the randomization protocol. We plotted the distribution of gift incentives in these villages and confirmed it was non-uniform before excluding them. We modeled the effect of gift amount on study participation, specifically consent and availability, with an OLS specification at the individual level with village and enumerator fixed effects and clustered standard errors by household. We conducted this analysis for both the first visit and all visits (first and revisit) combined. Consent was defined as providing informed consent for both the survey and sample collection. We estimate the effect of gift amounts using the following specification:

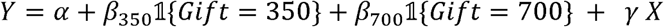

Where *Y* is a binary outcome indicating availability or consent, *β*_350_ and β_700_ are coefficients associated with the effect of receiving a KSh 350 gift and KSh 700 (compared to a baseline of KSh 200), and x is a vector including enumerator and village fixed effects. In a secondary analysis, we examined how timing impacted availability and consent by modeling the interaction between gift incentive amount and the percentage of a village that had already completed surveys at the time of an individual’s first household visit. Finally, we examine the association between incentives and infection rates.

We report the prevalence of cumulative infection overall and stratified by age-groups (0-5 years, 6-18 years, 19-60 years, >60 years), with bootstrapped 95% confidence intervals (CIs) (reps = 1000). Any infection was defined as a positive qPCR test (nasal swab) or antibody test by ELISA (DBS). We report infection outcomes as mean prevalence with 95% CIs. Additional analyses are described in SI. Additionally, we used a simple linear model to predict the number of consents under fixed incentive scenarios. We calculated the cost per visit, based on actual study costs, and estimated the cost per consent for each incentive amount.

Finally, we estimate the effect of non-response on our estimates of COVID-19 infection. To estimate the difference between COVID rates in initial and follow-up visits, we used the following specifications across individuals who tested:

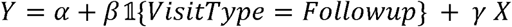

where *Y* is a binary outcome indicating a positive PCR or ELISA test for COVID-19, and *X* is either an empty vector or a vector containing week fixed effects. To estimate the difference between COVID rates by incentive, we used the following specifications across individuals who tested:

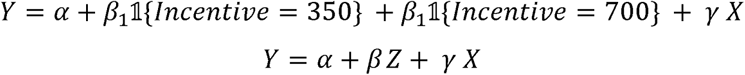

Where *Y* and *X* are as above and *Z* is the amount of compensation (in Kenyan shillings).

### Ethics

This study protocol was approved by the Scientific and Ethics Review Unit at Kenya Medical Research Institute (SERU Number 4111) and Committee for the Protection of Human Subjects at the University of California, Berkeley (2020-08-13592).

## Data Availability

All data produced in the present study are available upon reasonable request to the authors

## Acknowledgements

We thank the field staff and the communities involved in this study. YSC was supported by the Stanford University King Center on Global Development.

