## Supplemental Information for "Randomized incentives to increase participation in COVID testing in rural Kenya"

**Additional methods:**

*Sample processing*

Master mix reactions containing UltraPlex 1-Step ToughMix ROX (4X) master mix, Primer/Probe mix for N1, N2 and RNAse P assays were prepared. Five microlitres of RNA template sample and 15 microlitres of master mix were added to each well for each of the assays. Mixtures were dispensed in 96-well plates (MicroAmp™ Fast Optical 96-well reaction Plate 0.1 mL, Applied Biosystems) and sealed with optical film (MicroAmp™Optical Adhesive Film, Applied Biosystems). A positive control (nCoV-2 positive) and a no template control (negative control NTC) were included on each plate. Reactions were heated to 50^o^C for 10 minutes for reverse transcription, denatured in 95^o^C for 3 minutes, and then 45 cycles of amplification were carried in 95^o^C for 3 seconds and 55^o^C for 30 seconds. All no template controls were negative across plates.

DBS were eluted using 120 µL PBS + 0.05% Tween20 in 2 ml microcentrifuge tubes, vortexed thoroughly then incubated overnight at 4°C. The next day, samples were centrifuged for 2 minutes at 10,500 x g and the eluent was transferred to a clean microcentrifuge tube. 50 µl of negative and positive kit control and 100 µl of sample were added to the ELISA plate and incubated at 37 °C for 30 minutes. The plate was then washed 10 times with a diluted wash buffer provided by the kit. 100 µl of HRP-Conjugate was then added to each well except the blank well then incubated at 37 °C for 30 minutes. This was followed by washing the plate 10 times using a diluted wash buffer. After washing, 50 µl of chromogen Solution A and then 50 µl of Chromogen Solution B were added to each well then incubated at 37 °C in darkness (covered with aluminum foil) for 15 minutes. Finally, 50 µl of stop solution was added to each well and absorbance read at 450 nm. The Wantai SARS-CoV-2 Ab ELISA assay results were calculated as follows: If the ratio of absorbance to cut-off was <0.9 the results were interpreted as negative (no SARS-CoV-2 antibodies). If the ratio of absorbance to cut-off was ≥ 1.1 it was positive (have SARS-CoV-2 antibodies). If the ratio of absorbance to cut-off was between 0.9 and 1.1, the results were reported as equivocal, and the sample was re-tested.^1^

*Statistical analysis*

We used multivariable regression to model factors associated with risk of infection and report prevalence ratios using modified Poisson regression with robust standard errors. Models were stratified by age categories, due to differences in relevant factors and data collected by age groups. Covariates were included if they had a prevalence of at least 5%, were associated with the outcome based on a likelihood ratio test p-value <0.2, and if there were at least 10 events per covariate. We considered the following covariates for inclusion occupation category (agriculture/construction/labor, education, service, healthcare, government, student, retired); self-reported COVID risk reduction practices with 7-day recall (e.g., masking); self-reported exposure risks with 7-day recall (e.g., going to the market, travel); and respondent sex. A full list of covariates considered is available in [SI Table 12](https://docs.google.com/document/d/1oV-ZnNfy3rODOhxN3BTpQRcQ0kogGOrmk0xxPqMYpS4/edit?usp=sharing). In a post-hoc analysis, we used Fisher’s exact test to compare whether self reported symptoms were associated with positive/negative qPCR results.

**SI Table 1. Timing Analysis**

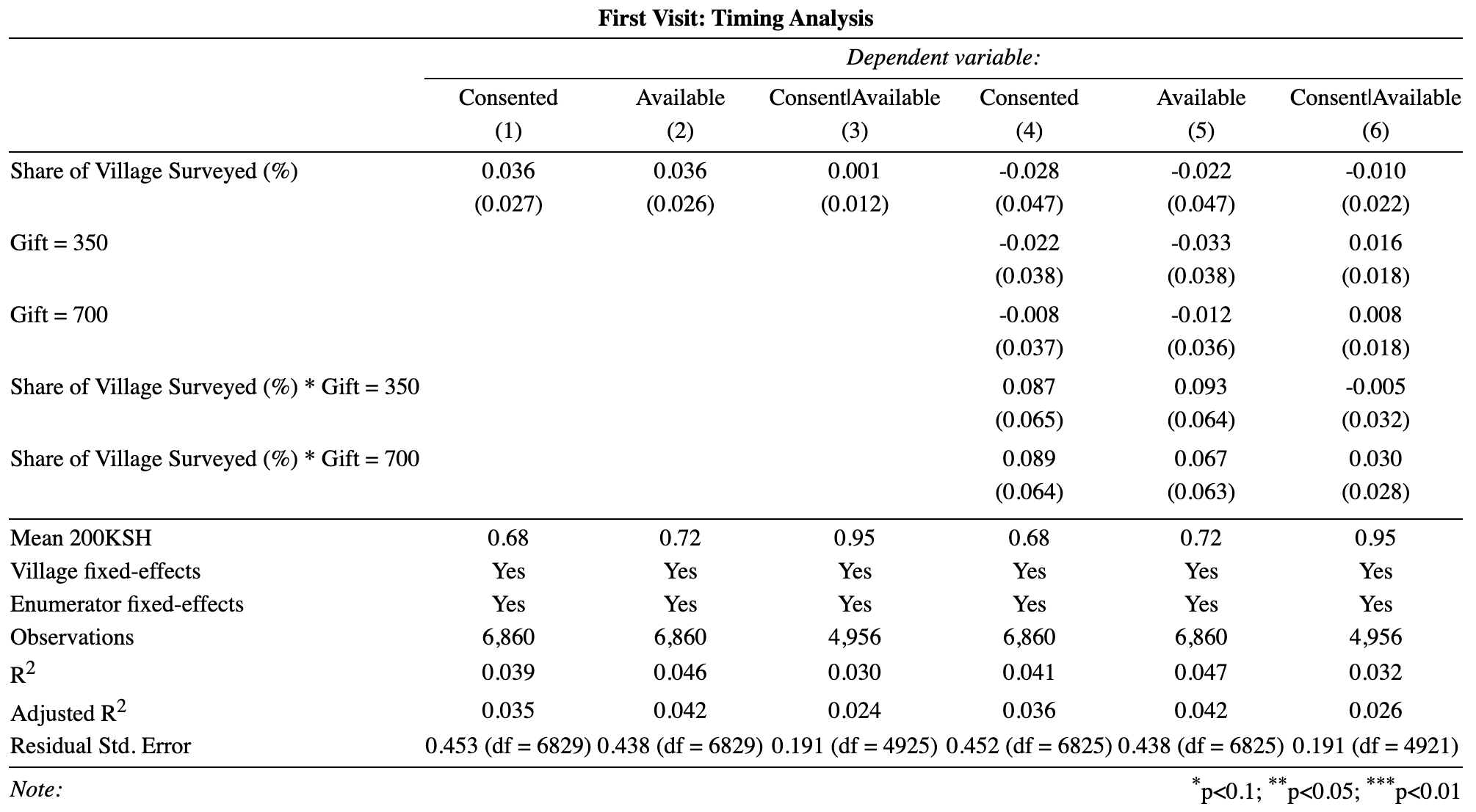

**SI Table 2.** Effect of gift amount on participation across all visits, stratified by county

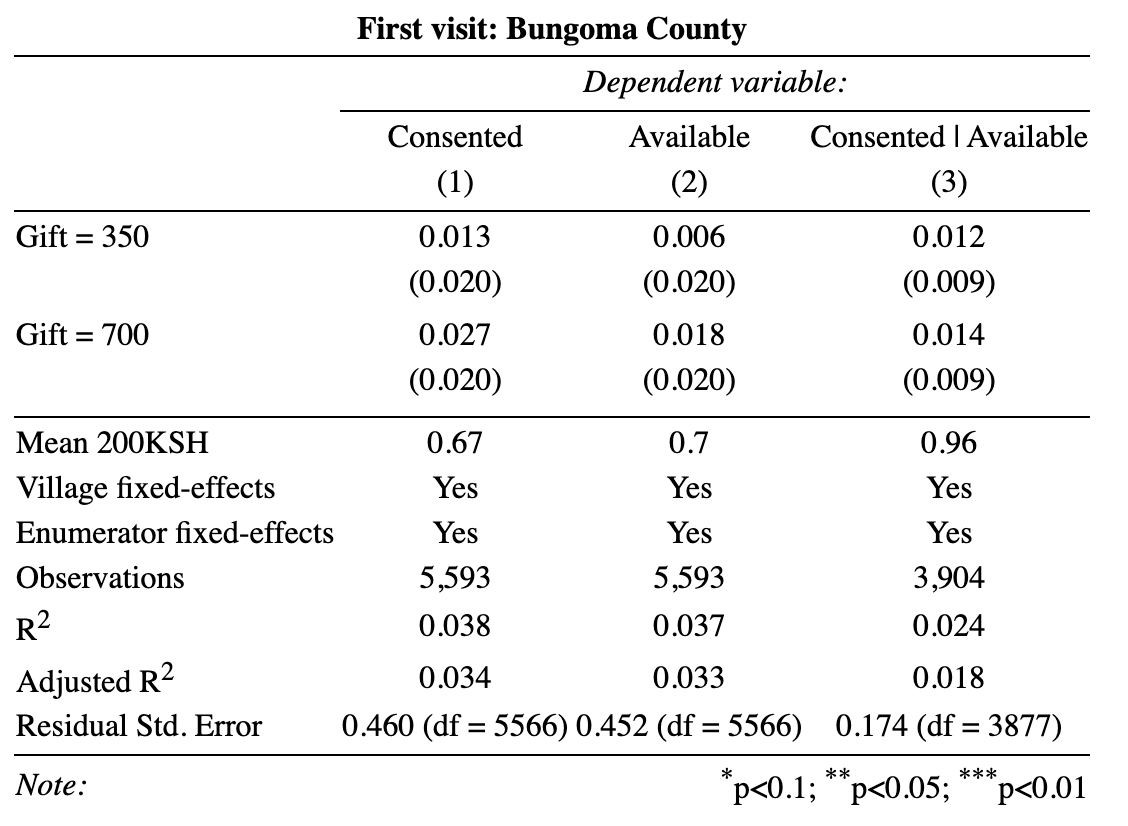

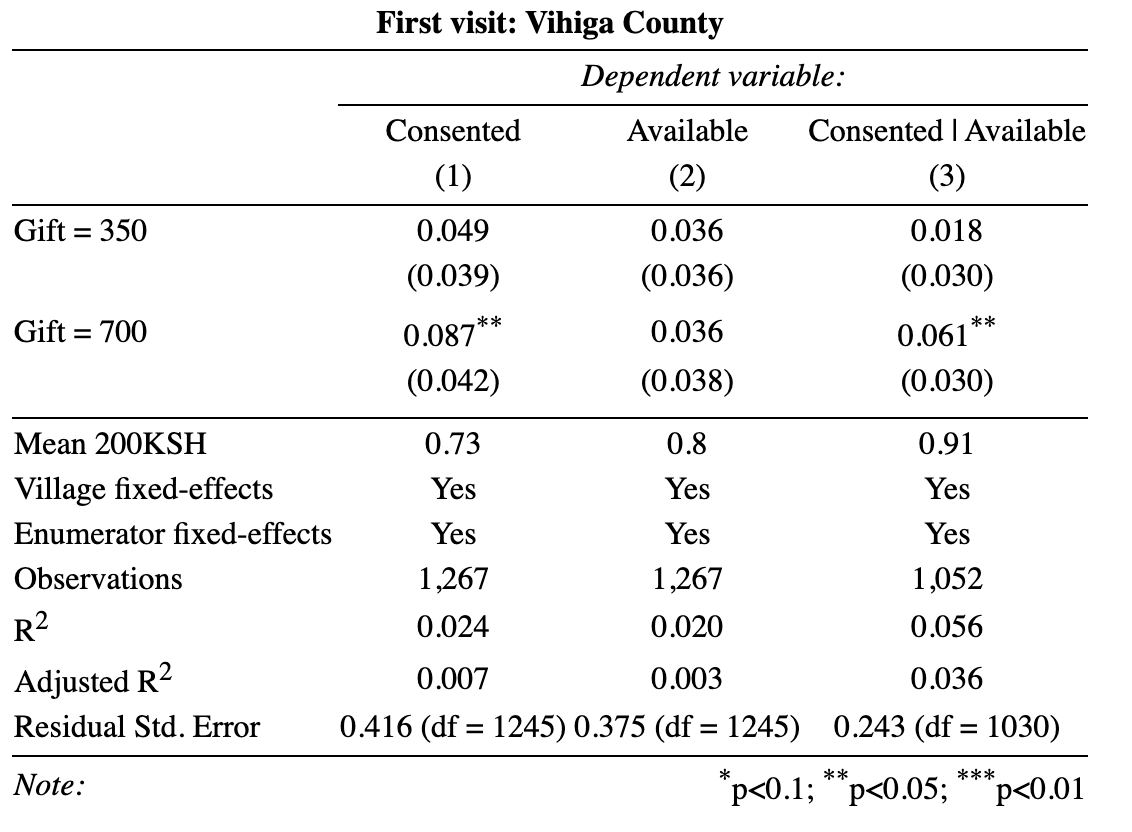

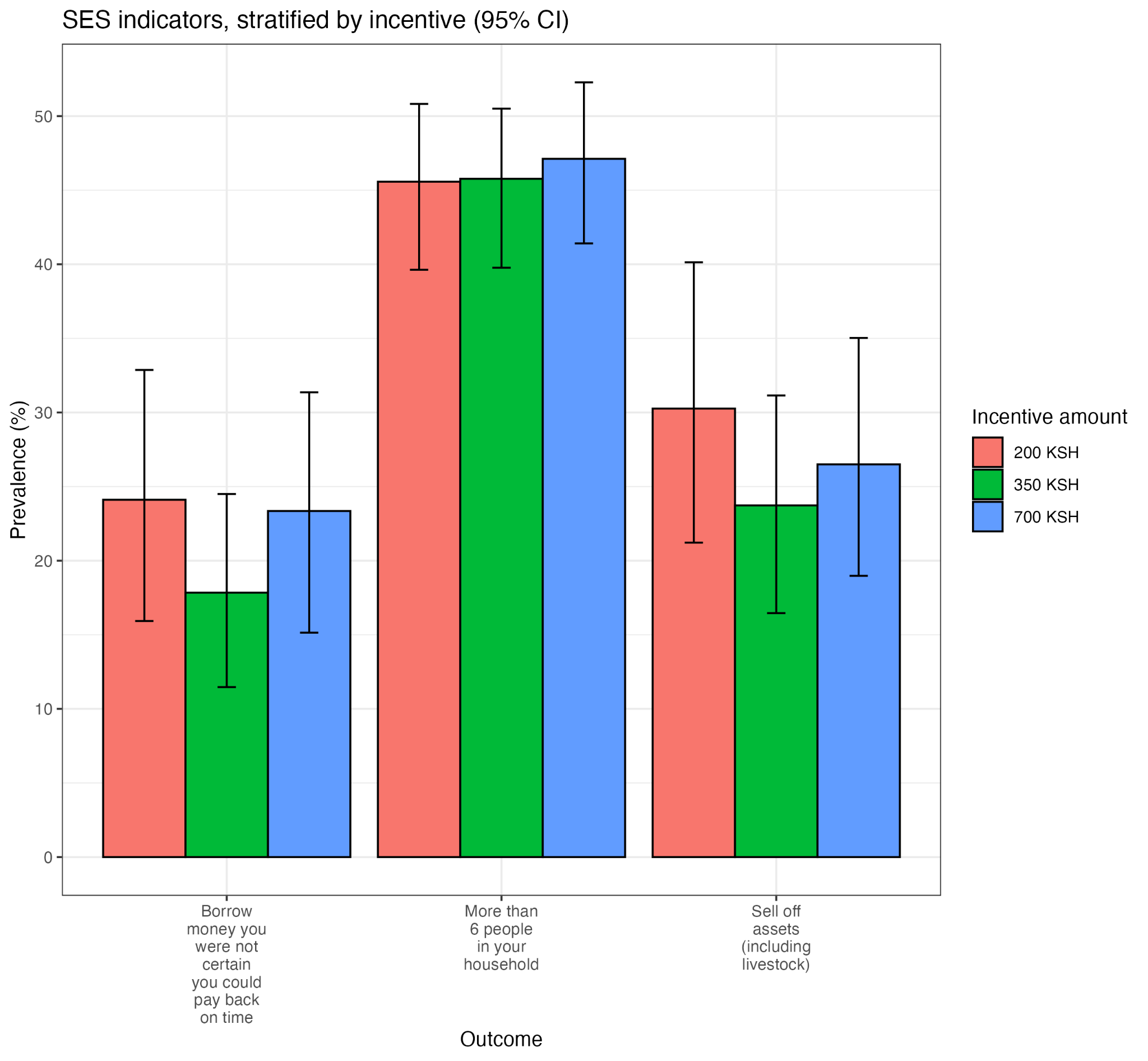

**SI Figure 1.** Indicators of financial status and coping among individual participants who consented, stratified by gift incentive

**SI Table 6.** Sex-stratified cumulative infection, seropositivity (ELISA), and current infection (qPCR)

|  | **Female** | | **Male** | |
| --- | --- | --- | --- | --- |
| **Outcome** | **n** | **Prevalence, % (95% CI)** | **n** | **Prevalence, % (95% CI)** |
| Cumulative infection | 3006 | 9.3 (8.1, 10.6) | 2380 | 8.6 (7.4, 10.0) |
| ELISA | 3033 | 8.9 (7.7, 10.1) | 2390 | 8.2 (7.0, 9.5) |
| qPCR | 3123 | 0.3 (0.1, 0.5) | 2494 | 0.4 (0.1, 0.6) |

**SI Table 7.** Age-stratified cumulative infection, seropositivity (ELISA), and current infection (qPCR)

|  | **0-5 years** | | **6-17 years** | | **18-60 years** | | **>60 years** | |
| --- | --- | --- | --- | --- | --- | --- | --- | --- |
| **Outcome** | **n** | **Prevalence, % (95% CI)** | **n** | **Prevalence, % (95% CI)** | **n** | **Prevalence, % (95% CI)** | **n** | **Prevalence, % (95% CI)** |
| Cumulative infection | 934 | 4.2  (2.9, 5.5) | 2279 | 8.8  (7.6, 10.2) | 1819 | 12.2  (10.7, 13.8) | 354 | 6.5  (4.0, 9.5) |
| ELISA | 942 | 4.0  (2.8, 5.3) | 2286 | 8.6  (7.4, 9.9) | 1838 | 11.6  (10.2, 13.1) | 357 | 5.3  (2.9, 8.1) |
| qPCR | 960 | 0.1  (0.0, 0.3) | 2389 | 0.2  (0.0, 0.4) | 1904 | 0.5  (0.2, 0.8) | 364 | 1.1  (0.3, 2.3) |

**SI Table 8.** County-stratified cumulative infection, seropositivity (ELISA), and current infection (qPCR)

|  | **Bungoma County** | | **Vihiga County** | |
| --- | --- | --- | --- | --- |
| **Outcome** | **n** | **Prevalence, % (95% CI)** | **n** | **Prevalence, % (95% CI)** |
| Cumulative infection | 4371 | 7.2 (6.3, 8.2) | 1015 | 16.7 (14.0, 19.6) |
| ELISA | 4406 | 6.9 (6.0, 7.8) | 1017 | 16.0 (13.3, 18.7) |
| qPCR | 4533 | 0.3 (0.1, 0.4) | 1084 | 0.6 (0.2, 1.2) |

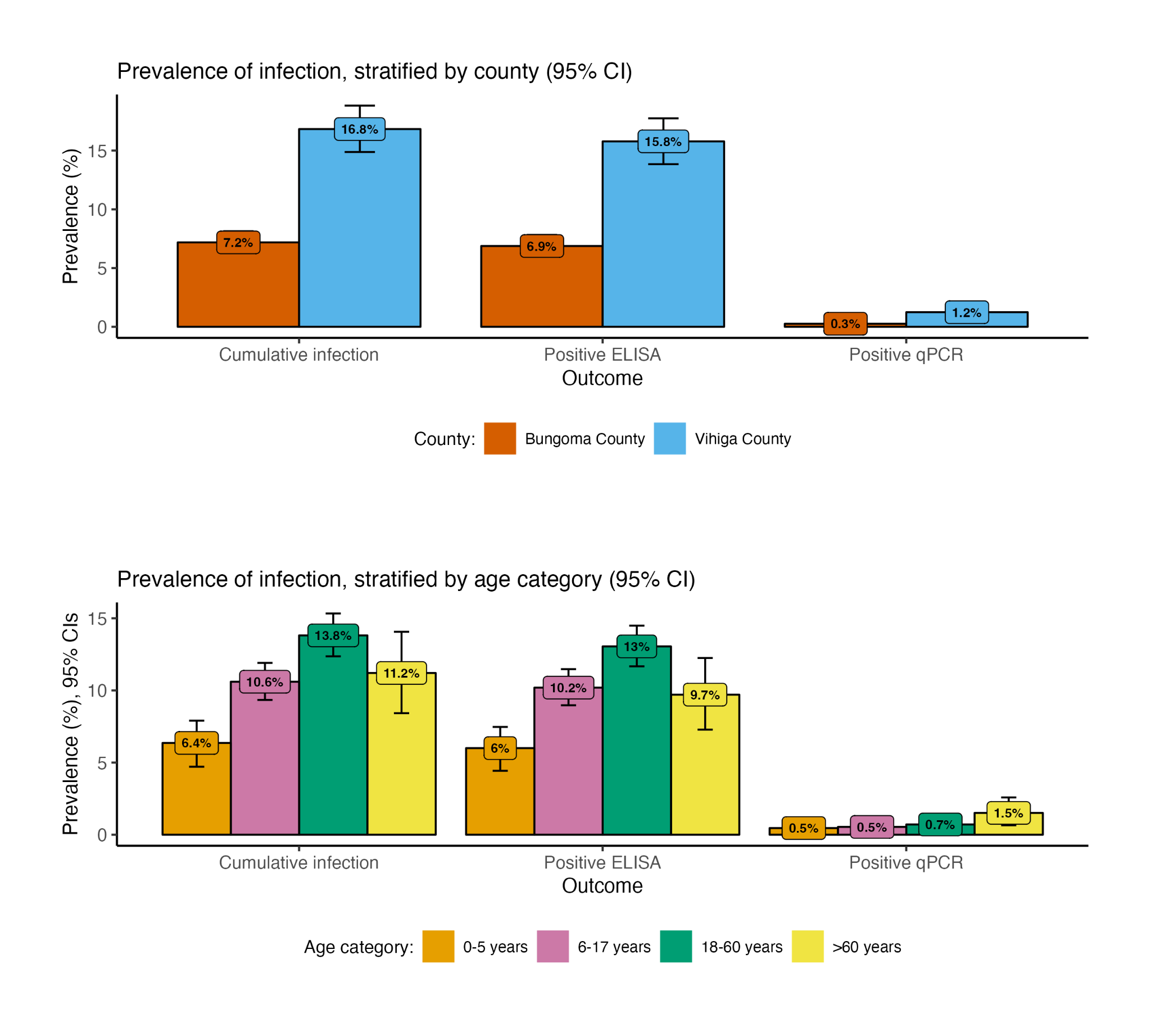

**SI Figure 2.** Prevalence of cumulative infection, seropositivity (ELISA), and current infection, stratified by county and age

**SI Table 9.** Self-reported symptoms with 7-day recall and COVID status via qPCR

|  | **COVID negative**  **(n=5,598)**  Mean (SD) | **COVID positive**  **(n=19)**  Mean (SD) | Fisher’s exact test  p-value |
| --- | --- | --- | --- |
| Cough | 0.13 (0.34) | 0.21 (0.42) | 0.32 |
| Headache | 0.13 (0.33) | 0.21 (0.42) | 0.27 |
| Congestion or runny nose | 0.10 (0.30) | 0.11 (0.32) | 0.95 |
| Fever | 0.07 (0.25) | 0.11 (0.32) | 0.501 |
| Chills | 0.04 (0.19) | 0.16 (0.37) | **0.006** |
| Unusual muscle pains | 0.03 (0.17) | 0.16 (0.37) | **0.002** |
| Unusual fatigue | 0.03 (0.16) | 0.11 (0.32) | **0.027** |
| Diarrhea | 0.02 (0.15) | 0.00 (0.00) | 0.49 |
| Difficulty breathing | 0.02 (0.15) | 0.00 (0.00) | 0.497 |
| Repeated shaking with chills | 0.02 (0.15) | 0.21 (0.42) | **<0.001** |
| Unusual pain or pressure in the chest | 0.02 (0.15) | 0.11 (0.32) | **0.018** |
| Loss of appetite | 0.02 (0.15) | 0.16 (0.37) | **<0.001** |
| Skin rash | 0.02 (0.13) | 0.00 (0.00) | 0.551 |
| Nausea or vomiting | 0.01 (0.12) | 0.05 (0.23) | 0.171 |
| Sore throat | 0.01 (0.10) | 0.00 (0.00) | 0.653 |
| Loss of smell/taste | 0.01 (0.09) | 0.00 (0.00) | 0.679 |
| Unexplained bruising | 0.01 (0.09) | 0.00 (0.00) | 0.701 |
| New confusion | 0.01 (0.07) | 0.00 (0.00) | 0.757 |

*Fewer than 1% of respondents reported inability to wake or stay awake in the prior 7 days

*Knowledge and risk perceptions*

19.9% (530/2667) of adults reported feeling that they or a household member were at risk of infection, although 93.4% (2492/2667) reported that they would get the vaccine when it became available to them. They most commonly and correctly mentioned cough (68.4% of respondents), fever (52.2%), headache (43.9%), and difficulty breathing (37.4%) as symptoms of COVID-19. Sneezing was mentioned by 24.4% of respondents ([SI](https://docs.google.com/document/u/0/d/1oV-ZnNfy3rODOhxN3BTpQRcQ0kogGOrmk0xxPqMYpS4/edit) Table 10). When asked what they would do if they or a family member had symptoms of COVID, 90.0% (2400/2667) said they would seek medical care at a health clinic or hospital, 4.9% (131/2667) would stay at home and isolate, and 4.3% (116/2667) would get tested for COVID-19. Other responses mentioned by more than 2% of respondents included calling or texting the Ministry of Health (3.0%, 81/2667), seeking care from a pharmacy (2.2%, 60/2667), and wearing a face mask or covering (2.0%, 54/2667) ([SI](https://docs.google.com/document/u/0/d/1oV-ZnNfy3rODOhxN3BTpQRcQ0kogGOrmk0xxPqMYpS4/edit) Table 11).

**SI Table 10.** Knowledge: symptoms of COVID as mentioned by adults, unprompted list

|  | Overall  n (%) |
| --- | --- |
|  | n=2,667 |
| Cough | 1825 (68.4) |
| Fever | 1393 (52.2) |
| Headache | 1172 (43.9) |
| Difficulty breathing | 997 (37.4) |
| Sneezing | 650 (24.4) |
| Congestion or runny nose | 273 (10.2) |
| Chills | 212 (7.9) |
| Sweating | 162 (6.1) |
| Sore throat | 158 (5.9) |
| Unusual fatigue | 154 (5.8) |
| Unusual muscle pains or body aches | 126 (4.7) |
| Unusual pain or pressure in the chest | 93 (3.5) |
| Diarrhea | 91 (3.4) |
| Nausea or vomiting | 68 (2.5) |
| Repeated shaking with chills | 36 (1.3) |
| Loss of smell/taste | 35 (1.3) |
| Stomach pain | 33 (1.2) |
| Loss of appetite | 28 (1.0) |
| New confusion | 6 (0.2) |
| Gray or discolored lips or face | 5 (0.2) |
| Inability to wake or stay awake | 4 (0.1) |

**SI Table 11.** Covid response: What would you do if you/family member had symptoms of COVID?

|  | Overall  n (%) |
| --- | --- |
|  | n=2,667 |
| Seek medical care at a health clinic or hospital | 2400 (90.0) |
| Stay at home/isolate | 131 (4.9) |
| Get tested for COVID-19/coronavirus | 116 (4.3) |
| Call/text Ministry of Health | 81 (3.0) |
| Seek care from a pharmacy | 60 (2.2) |
| Wear a face mask or covering | 54 (2.0) |
| Wash hands more frequently | 50 (1.9) |
| Physical/social distance | 49 (1.8) |
| Do nothing | 48 (1.8) |
| Don't know | 29 (1.1) |
| Visit a traditional healer | 8 (0.3) |
| Pray | 5 (0.2) |

*Risk-associated factors*

No factors were associated with a positive ELISA result among adults at a p=0.05 significance level. However, staying home more in the prior 7 days (adjusted PR: 0.79 (0.59, 1.06; p=0.11)) suggested lower risk, and working in service occupations (aPR: 1.36 (0.95, 1.95; p=0.09)) and more than 3 visits to a market or food store by the respondent or other household member in the prior 7 days (aPR: 1.26 (0.99, 1.60; p=0.06) suggested increased risk. Avoiding shaking hands in the prior 7 days to reduce COVID risk (aPR: 0.96 (0.73, 1.26; p=0.77) met covariate screening criteria but was not associated with the outcome in the full model ([SI](https://docs.google.com/document/d/1oV-ZnNfy3rODOhxN3BTpQRcQ0kogGOrmk0xxPqMYpS4/edit) Table 13).

There was insufficient data to model factors associated with qPCR or results restricted to ages 60+.

**Occupation categories**

The survey included occupation options, but respondents were also allowed to describe their occupation as a free response question. The following is the categorization of all recorded occupation responses.

*Agricultural/construction/labor:* "Agriculture, forestry, or fishing - Subsistence (personal/household consumption)", "Agriculture, forestry, or fishing - Commercial (goods sold or traded)", "Construction, carpentry, masonry, fabricators", "Electricity, gas, steam, or air conditioning supply", "Manufacturing", "Mining/quarrying", "Water supply, sewage, or waste management", "Truckloading", "Watchman/Caretaker", "Mechanic, repair of motor vehicles and motorcycles", "Weaving", "Washing and cleaning the environment.", "Washing", "Transporting stones", "Timber making", "Stone digging", "Spliting firewood", "Spliiting of firewood", "Smearing the house", "Poshomill", "Plumbing", "Plant operator", "Mason", "Looking after cattle", "Looking after animals", "Herding of cows", "Herding cattle", "Gardening for other people to get paid", "Gardening for other people", "Gardening and farming", "Fetching water", "Fetching firewood", "Digging latrine", "Digging borehole", "Cutting grass for the animal", "Construction", "Collection of sand from the river to sell", "Collecting charcoal", "Charcoal burning.", "Cattle herding", "Carpentry", "Carpenter", "Cane cutter", "Burning charcoal", "Burning charcoal", "Bricks making", "Brick making"

*Education:* "Education", "Research", "Teacher by profession", "Teacher"

*Service industry:* "Financial and insurance activities", "Real estate", "Salonist", "Tailor", "Wholesale and retail trade", "Tailoring and business", "Solar marketing", "Small business", "Shoppkepper", "Shopkeeper", "Shop keeper", "Shoe making", "Selling sukuma wiki", "Selling sugarcane", "Selling sambuza", "Selling ripe bananas", "Selling napper grass", "Selling mutumba", "Selling milk and groceries", "Selling maize", "Selling groceries", "Selling firewood", "Selling eggs", "Selling business (locally brewed alcohol)", "Selling omena", "Selling omega and fish", "Selling firewood", "Sell brew", "Ripe banana and vegetables", "Pots seller", "Operating M-pesa shop", "Mama mboga", "Maid", "M-pesa agent", "Loundry", "Local brew", "Laundry", "Juakali", "Jua kali", "Housework", "Houseboy", "House-helper", "House work", "House help to some old mama", "Guide in the village", "Guide", "Fabric", "Cook", "Cleaning home", "Chef at the finerals", "Buying and selling cattle", "Business woman", "Business", "Brewing alcohol", "Bodaboda rider", "Baby sitting"

*Healthcare:* "Healthcare", "Social work activities", "Traditional herbalist", "Community health voluntere", "Community health volunteer", "Community Health volunteer", "Commnity health volunteer", "CHV", "Birth companion"

*Government:* "Government employee"

*Student:* "Student still", "Student", "School pupil", "School", "Pupil"

*Retired:* "Retired civil servant", "Retired civil servants", "Old age doesn't do productivity", "Housewife", "House chores Cleanliness of the compound", "House chores", "Home activities", "Elderly monthly coupons"

**SI Table 12. Covariates assessed for inclusion in model**

| Category | Variables |
| --- | --- |
| *Demographics* | Sex, occupation category (see above occupation categorization), number of household members (6 or fewer vs more than 6) |
| *Self-reported behaviors & exposures* | *In the past 7 days, self-reporting the following measures:* wearing a facemask or covering, cleaning surfaces, staying at home more, avoid touching face, avoid shaking hands, handwashing, avoiding hospitals/clinics, avoiding large gatherings, encouraging children to play at home, reducing contact with other people/social distancing, staying aware of the latest information on the outbreak, attending church/mosque/prayer gathering, meeting with a merry go round savings group, attending a funeral or burial, visiting a bar or restaurant, number of non-household contacts, number of visits (3 or fewer vs more than 3) (self or other household member) to market or food store,  *In the past 30 days:* travel (self or other household member) to or from a city or border town, visitors (from outside household) from a city or border town, close contact with anyone positive for COVID-19  *Yesterday:* number of hours spent outside the respondent’s compound, number of handwashing events with soap or hand sanitizer  *Other:* Materials used for handwashing the last time the respondent washed hands |

**SI Table 13. Factors associated with positive ELISA, ages 18+**

|  | **PR** | **95% CI** | **p-value** |
| --- | --- | --- | --- |
| In the past 7 days, stayed home more | 0.79 | 0.59, 1.06 | 0.11 |
| In the past 7 days, avoided shaking hands | 0.96 | 0.73, 1.26 | 0.77 |
| Works in a service industry job | 1.36 | 0.95, 1.94 | 0.09 |
| In the past 7 days, >3 visits to a market of food store by self or a household member | 1.26 | 0.99, 1.60 | 0.06 |

**SI Table 16. Prevalence of infection, stratified by gift incentive**

|  | **200** | | **350** | | **700** | |
| --- | --- | --- | --- | --- | --- | --- |
| **Outcome** | **n** | **Prevalence, % (95% CI)** | **n** | **Prevalence, % (95% CI)** | **n** | **Prevalence, % (95% CI)** |
| Cumulative infection | 1675 | 7.6  (6.3, 8.9) | 1868 | 9.0  (7.4, 10.7) | 1843 | 10.2  (8.5, 12.0) |
| qPCR | 1749 | 0.5  (0.2, 0.8) | 1946 | 0.3  (0.0, 0.6) | 1922 | 0.3  (0.1, 0.6) |
| ELISA | 1686 | 7.1  (5.8, 8.4) | 1880 | 8.7  (7.2, 10.5) | 1857 | 9.8  (8.1, 11.6) |

**SI Table 17. Infection Rates by Visit Type, by type of COVID test**

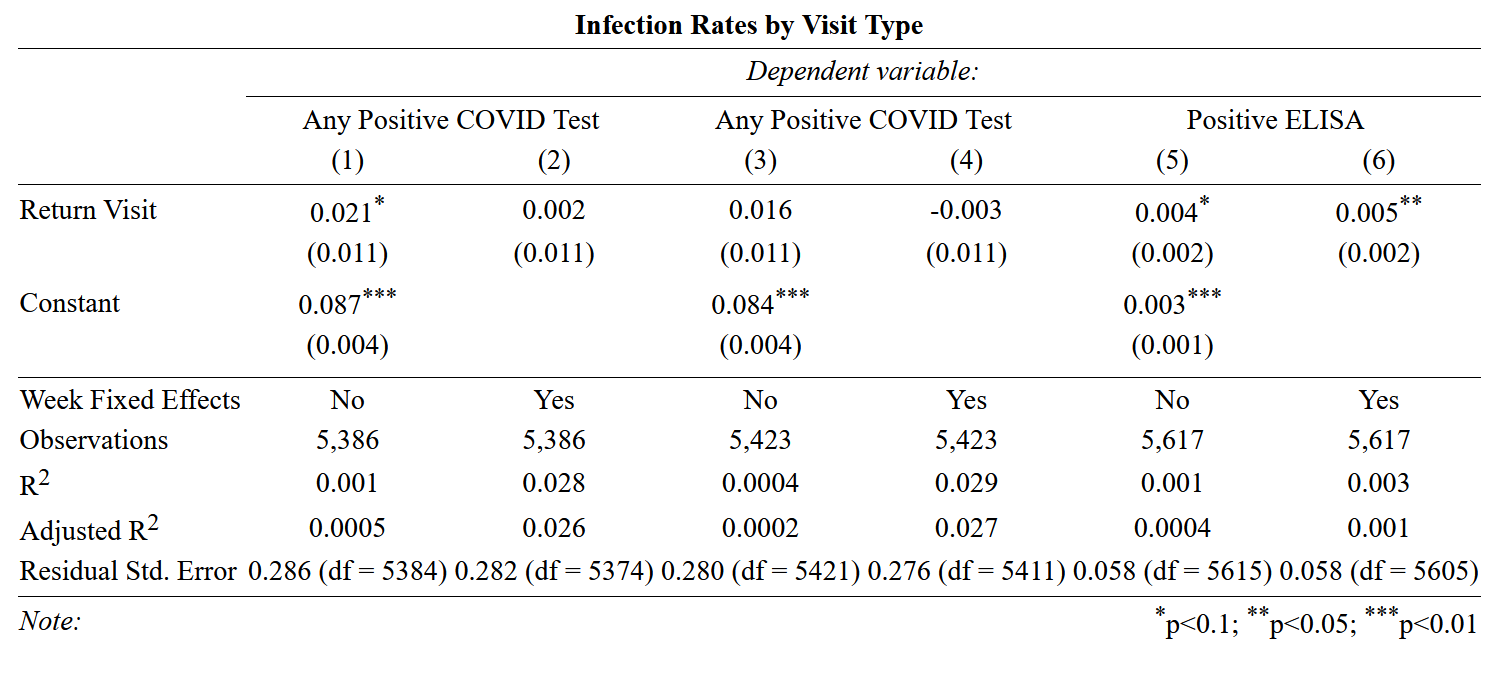
